# Substantial Impact of Post Vaccination Contacts on Cumulative Infections during Viral Epidemics

**DOI:** 10.1101/2020.12.19.20248554

**Authors:** Nash D. Rochman, Yuri I. Wolf, Eugene V. Koonin

## Abstract

**Background:** The start of 2021 will be marked by a global vaccination campaign against the novel coronavirus SARS-CoV-2. Formulating an optimal distribution strategy under social and economic constraints is challenging. Optimal distribution is additionally constrained by the potential emergence of vaccine resistance. Analogous to chronic low-dose antibiotic exposure, recently inoculated individuals who are not yet immune play an outsized role in the emergence of resistance. Classical epidemiological modelling is well suited to explore how the behavior of the inoculated population impacts the total number of infections over the entirety of an epidemic.

**Methods:** A deterministic model of epidemic evolution is analyzed, with 7 compartments defined by their relationship to the emergence of vaccine-resistant mutants and representing three susceptible populations, three infected populations, and one recovered population. This minimally computationally intensive design enables simulation of epidemics across a broad parameter space. The results are used to identify conditions minimizing the cumulative number of infections.

**Results:** When an escape variant is only modestly less infectious than the originating strain within a naïve population, there exists an optimal rate of vaccine distribution. Exceeding this rate increases the cumulative number of infections due to vaccine escape. Analysis of the model also demonstrates that inoculated individuals play a major role in the mitigation or exacerbation of vaccine-resistant outbreaks. Modulating the rate of host-host contact for the inoculated population by less than an order of magnitude can alter the cumulative number of infections by more than 20%.

**Conclusions:** Mathematical modeling shows that optimization of the vaccination rate and limiting post-vaccination contacts can affect the course of an epidemic. Given the relatively short window between inoculation and the acquisition of immunity, these results might merit consideration for an immediate, practical public health response.

## Introduction

The emergence of the novel Severe Acute Respiratory Syndrome Coronavirus 2 (SARS-CoV-2) responsible for the Covid-19 pandemic motivated dramatic public health intervention including recommendations for isolation and quarantine throughout most of 2020^1^. The beginning of 2021 will be marked by widespread vaccine distribution. Optimizing distribution is challenging and subject to a myriad of social and economic constraints^2-4^. The potential emergence of vaccine-resistant variants of the virus^5,6^ introduces additional complications. Vaccination applies new selective pressures which can lead to diverse intermediate outcomes even under conditions admitting eventual pathogen eradication^7-13^. The larger the size of the vaccinated population, the greater the pressure towards escape of vaccine-resistant variants.

Escape variants emerge within individual hosts after infection with the originating strain. Naïve, unvaccinated, hosts are more easily infected than vaccinated hosts but mutations conferring resistance are unlikely to provide a selective advantage in the naïve background. Thus, naïve hosts are likely to shed escape variants at very low, likely, negligible rates. The reverse is true for vaccinated hosts. Recently vaccinated, inoculated, hosts that are not yet immune remain highly susceptible to infection with the originating strain, and in these hosts, mutations conferring resistance are more likely to provide a selective advantage. As a result, a substantial fraction or even most of the virus shed by such hosts will be resistant mutants. This situation is analogous to the administration of a low-dose antibiotic regime^14,15^. In both cases, the pathogen is introduced to a susceptible host and subject to elevated selective pressure towards the emergence of resistant (escape) variants.

We sought to establish the constraints imposed by virus escape on optimal vaccine distribution and the role played by the small, but critical, population of inoculated hosts. To this end, we constructed an epidemiological compartment model to simulate vaccination campaigns over a broad parameter regime. This minimally computationally intensive approach enabled us to simulate many possible scenarios for epidemic evolution, in order to determine the optimal vaccination strategy for each condition.

## Methods

We divided the population into 7 compartments (Fig. 1A). Three compartments are susceptible to infection by either the originating strain or escape mutants: Naïve (*N*, unvaccinated and fully susceptible to the originating strain and escape mutants), Inoculated (*I*, recently vaccinated and still partially susceptible to the originating strain, and fully susceptible to escape mutants), and Vaccinated (*V*, minimally susceptible to the originating strain, but fully susceptible to escape mutants). Two compartments represent ongoing infection with the originating strain and are distinguished by the host’s previous compartment: Infected-Naïve (*F*) and Infected-Inoculated (*M*). The third infected compartment represents infection by an escape variant, Infected-Escape (*E*). The remaining compartment, Recovered (*R*), contains all hosts who were previously infected. Vaccination is represented by a reduction in susceptibility to infection with the originating strain. Naïve hosts are inoculated at rate *k*_*V*_. Inoculated hosts do not immediately acquire immunity and mature into the vaccinated compartment at rate *k*_*M*_. All infected hosts recover at rate *k*_*R*_.

**Figure 1.**
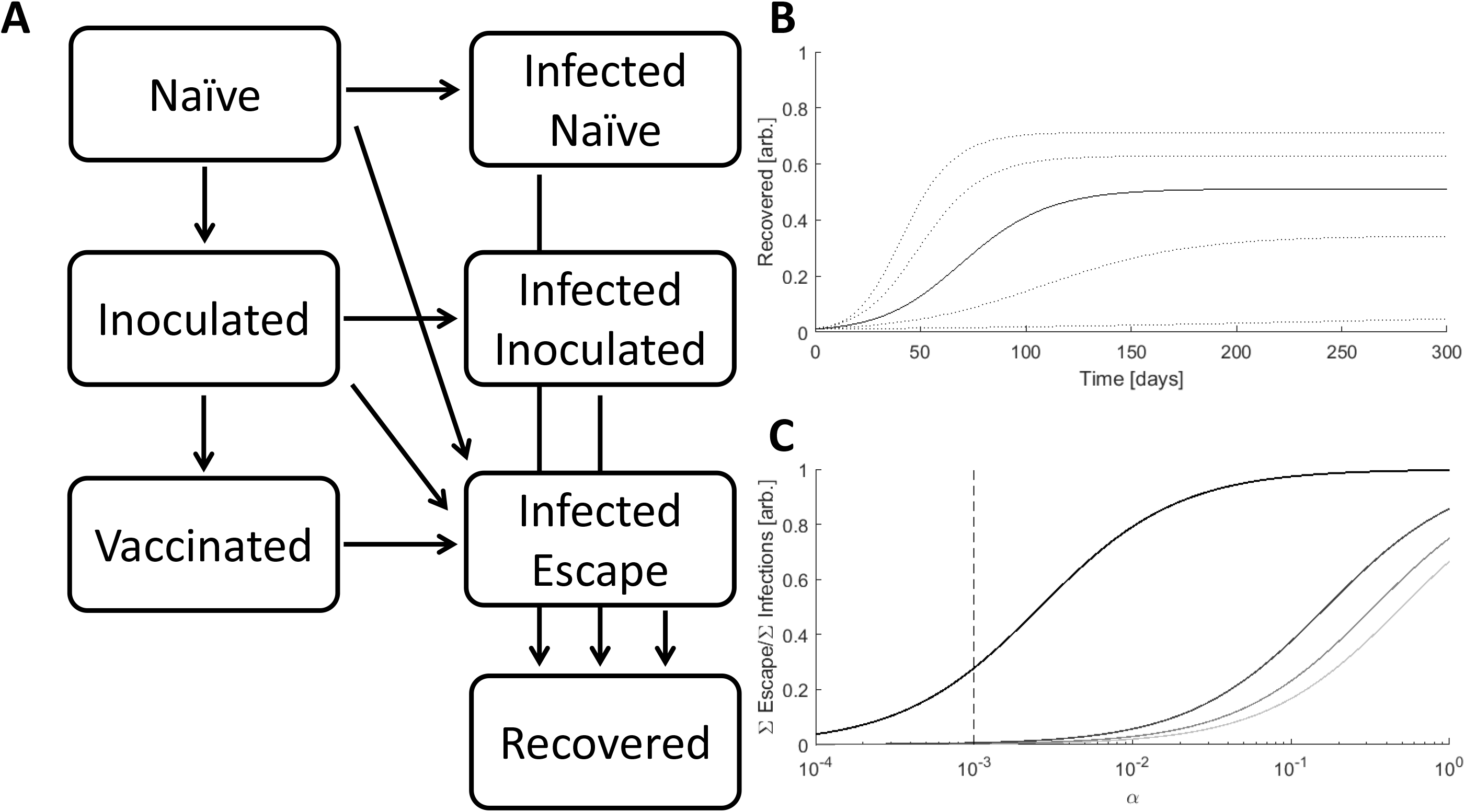
The Model. **A**. Schematic of the 7 compartment model with three susceptible, three infected, and one recovered compartments. **B**. Simulated epidemics for *k*_*I*_*=*[*0*.*15,0*.*175,0*.*2*(solid line),*0*.*225,0*.*25*], *α=0*.*001, β=0*.*01* **C**. The ratio of cumulative escape infections to all cumulative infections for an epidemic over a range of *α. β=*[*0*.*5,0*.*67,0*.*83,1*], darker color indicates higher value, *k*_*I*_*=0*.*2*. The dotted line specifies the benchmark value of *α=0*.*001*.

Within the timescale of the model, recovery is assumed to grant stable immunity, and any variation in population size due to birth/death is assumed to be negligible. It should be noted that, if recovery from the Infected-Naïve or Infected-Inoculated compartments does not confer immunity against escape infection, the key results in this work will have an even greater impact on the vaccination outcome. Hosts come into contact at rate *k*_*C*_. For simplicity, we assume that contact with an escape-infected host can only produce an escape infection. Also, vaccine efficacy is assumed to be perfect such that vaccinated hosts cannot be infected with the originating strain. The Inoculated-Infected compartment is assumed to represent a symmetric composition of escape and originating infections such that the total probability of a Naïve or Inoculated host being infected after contact with a Naïve-Infected or Inoculated-Infected host is the same.

Finally, we assume that the probability of escape-infection is the same for Naïve and Vaccinated hosts across all three types of infected-susceptible host interactions. This construction yields the following transition probability matrices for Naïve, Inoculated, and Vaccinated hosts:

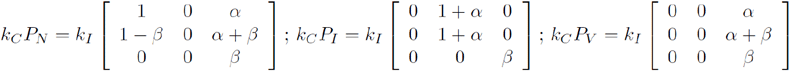

where *k*_*I*_ represents the rate of infection for Naïve, Infected-Naïve host interactions which is determined both by the contact rate *k*_*C*_ and the infectivity of the originating strain. Rows represent interaction with each of the infected compartments (Infected-Naïve, Infected-Inoculated, and Infected-Escape). Columns represent transitions to each of the infected compartments.

An escape mutant can emerge within an Infected-Naïve or Infected-Inoculated host. The parameter *α* represents the infectivity of the escape variant relative to the originating strain when a Naïve host interacts with an Infected-Naïve host. The parameter *β* represents the infectivity of an escape variant when a Naïve host interacts with an Infected-Escape host relative to the infectivity of the originating strain when a Naïve host interacts with an Infected-Naïve host. Informally, *α* reflects the ratio of escape variant to originating strain shed by Infected-Naïve hosts, whereas *β* reflects the fitness of an escape variant relative to the originating strain. Finally, we introduce the parameter *q* to represent the impact of varying the rate of host-host contact for Inoculated hosts relative to that for the other compartments. *q*>*1* represents increased contact, and *q*<*1* corresponds to decreased contact. This completes the model description and structures the differential equations:

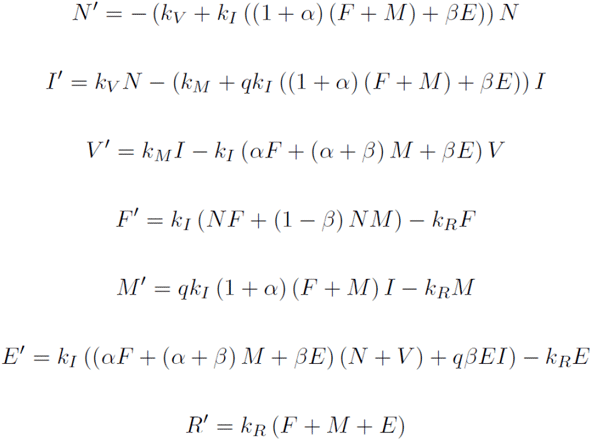

*k*_*R*_ = *k*_*M*_=*1*/*7* are fixed across all simulations representing a time to recovery and time between inoculation and the acquisition of immunity of one week. Reducing *k*_*I*_ would prolong the epidemic and reducing *k*_*M*_ would increase the size of the Inoculated compartment.

*k*_*I*_ is the principal determinant of epidemic magnitude and duration, with larger k_I_ leading to a greater cumulative number of infections over a shorter period of time (Fig. 1B). However, feedback between the size of the infected population and the rate of host-host contact as well as spatial structure can decouple these variables. Throughout this work, *k*_*I*_ is set to a benchmark value of 0.2 resulting in 50% of the population being infected over a period of approximately 4 months.

The values of *α* and *β* impact the size of the Infected-Escape compartment. Even in the absence of vaccination, large *α*/*β* results in the emergence of would-be resistant variants (Fig 1C). In all analyses in this work, *α* is fixed at the benchmark value of 0.001 resulting in a modest number of would-be resistant infections for *β* close to 1 in the absence of vaccination. Although a larger *α* would result in a greater total number of escape infections, the fraction of those infections attributable to contact with inoculated hosts would be smaller.

The solutions of the ODEs were obtained using the MATLAB ode45 method^16^. Epidemics are simulated until the size of the Recovery compartment at arbitrarily long times is approached. The principal quantity of interest is the cumulative number of infections. When *k*_*V*_ is selected to minimize this value, minima are found through explicit simulation over a range of rates. In the subsequent analysis, some values are expressed relative to the cumulative number of infections in the absence of vaccination, *R*_*Null*_∼*50%*.

## Results

In addition to the rate of vaccination, the outcome of a vaccination campaign depends on how far the epidemic has progressed before vaccination begins, which can be measured by the relative size of the Recovered compartment. The results also depend on *β*, informally, the fitness of an escape variant relative to the originating strain. We considered two values for *β* (0.01, low; 0.875, high) and varied both the start and rate of vaccination. When *β* is low, that is, the escape mutant is much less fit than the originating strain (Fig. 2A), vaccinating earlier and distributing the vaccine faster decreases the cumulative number of infections. If distribution is sufficiently prompt, the cumulative number of infections becomes negligible.

**Figure 2.**
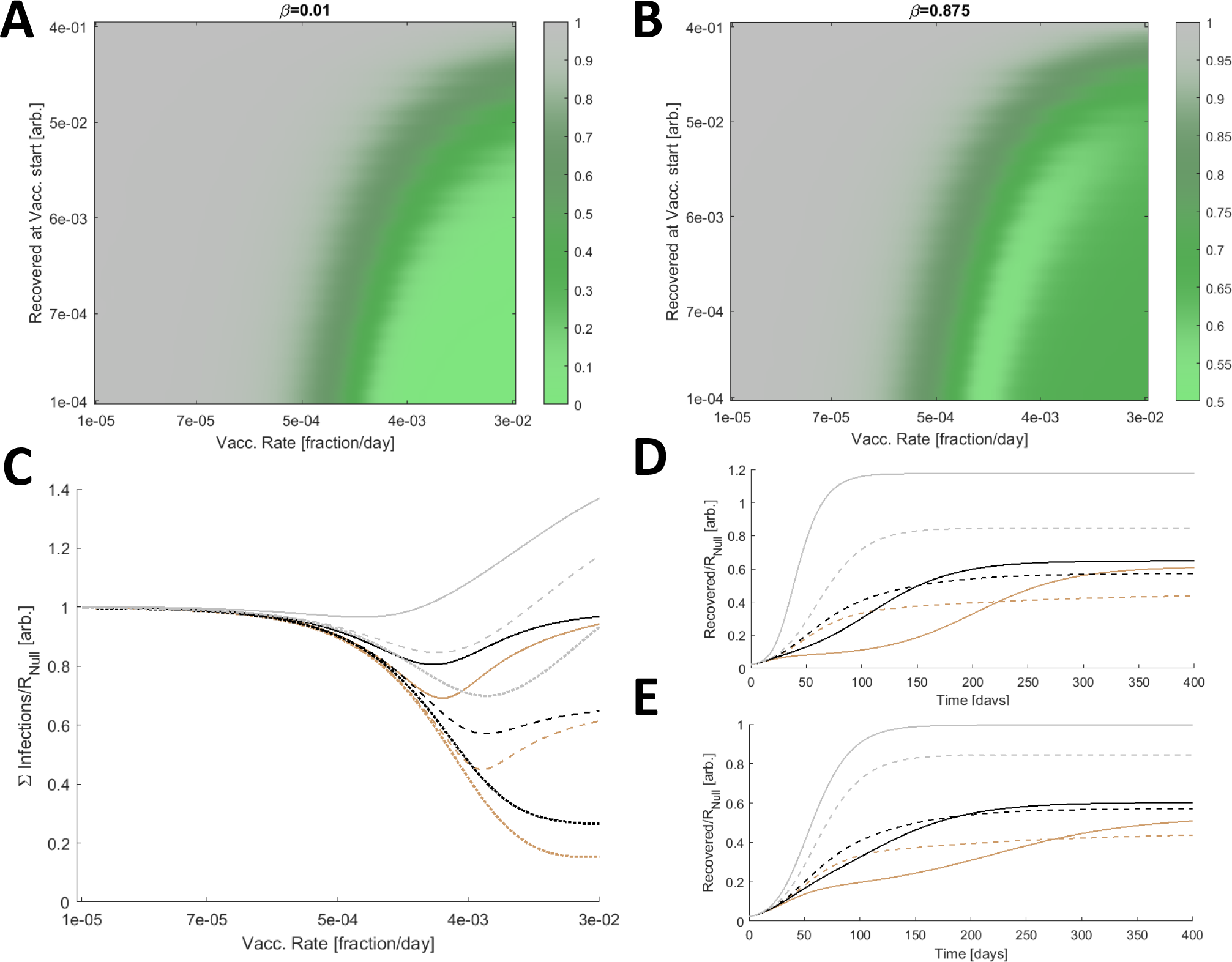
Optimal Vaccine Distribution. **A**. The cumulative number of infections relative to no vaccination, *R*_*Null*_, for a range of vaccine initiations and distribution rates. Here *β* is low, 0.01. **B**. The cumulative number of infections relative to no vaccination, *R*_*Null*_, for a range of vaccine initiations and distribution rates and a large *β* (0.875). **A/B**. 3840 values were computed for each panel and 4x by 4x bilinearly interpolated points are displayed. **C**. The cumulative number of infections relative to no vaccination, *R*_*Null*_, for *β=*[*0*.*75*(dotted),*0*.*875*(dashed),*1*(solid)] and three relative contact rates, *q=*[*0*.*2*(brown),*1*(black),*5*(gray)]. **D**. Simulated epidemics comparing a high fixed rate of vaccination, *k*_*V*_=*0*.*03* (solid) representing the inoculation of 3% of Naïve hosts per day, and the optimal vaccination rate for each condition (dashed). *β=0*.*875* is fixed and *q=*[*0*.*2*(brown),*1*(black),*5*(gray)]. **E**. Same as D. with a low fixed rate of vaccination, *k*_*V*_=*0*.*01*. **C/D/E**. The minima for the dashed lines in C correspond to the dashed lines in D/E.

However, the outcome substantially differs for high *β* (Fig. 2B). Due to the vaccine escape, at high rates of distribution, the cumulative number of infections remains large. Furthermore, there is an optimal rate of distribution such that exceeding this rate increases the cumulative number of infections. In this regime, the benefit of reducing the size of the population susceptible to infection by the originating strain is outweighed by the cost of increasing the selective pressure for the emergence of escape variants. In all subsequent analysis, the vaccination rate is varied but vaccination is fixed to begin when 1% of the population has recovered from infection.

Infections can be mitigated by reducing host-host contact. We sought to determine how perturbing the contact rates for hosts in the Inoculated compartment relative to that of all other compartments, *q*, affects the outcome. For *β* ranging between 0.75 and 1, we considered three relative contact rates, *q=*[*0*.*2,1,5*] (Fig. 2C). Increasing the rate of host-host contact only within this compartment has a significant impact on the cumulative number of infections. The optimal vaccination rate is also perturbed. Furthermore, if the optimal vaccination rate it exceeded, reducing *q* below 1 slows the accumulation of infections (Fig. 2D). The converse is true for increasing *q*, and the landscape is similar when vaccination falls below the optimal rate (Fig. 2E).

Having demonstrated how *q* perturbs the optimal vaccination rate and how reducing *q* below 1 can mitigate or delay the cumulative number of infections even if this rate is not met or, conversely, is exceeded, we sought to establish the impact of *q* on the cumulative number of infections across a wide range of *β* at the optimal vaccination rate for each condition (Fig. 3A). The cumulative number of infections is sensitive to *q* across the entire range of *β*. Varying *q* within an order of magnitude can substantially aid or hinder the vaccination campaign, and when *q*>>*1*, the optimal rate of vaccination is 0. Note that the maximum vaccination rate considered is *k*_*V*_=*0*.*03* representing the inoculation of 3% of Naïve hosts per day. This rate is sufficiently high that the cumulative number of infections at this rate (Fig. 3B) is similar or higher compared to the hypothetical case where the entire Naïve population is immediately inoculated (Fig. 3C). Some of these effects are not specific to the inoculated compartment. Increasing the rate of contact for any arbitrary subpopulation can increase the cumulative number of infections. We define the effective *k*_*I*_, *kIeff*, such that the cumulative number of infections for *k*_*I*_=*kIeff* and *q=1* is equal to the cumulative number of infections for *k*_*I*_=0.2 (the benchmark) and arbitrary *q*. Increasing *q* within an order of magnitude is equivalent to substantially increasing *k*_*I*_ for the entire population for the entirety of the epidemic (Fig. 3D).

**Figure 3.**
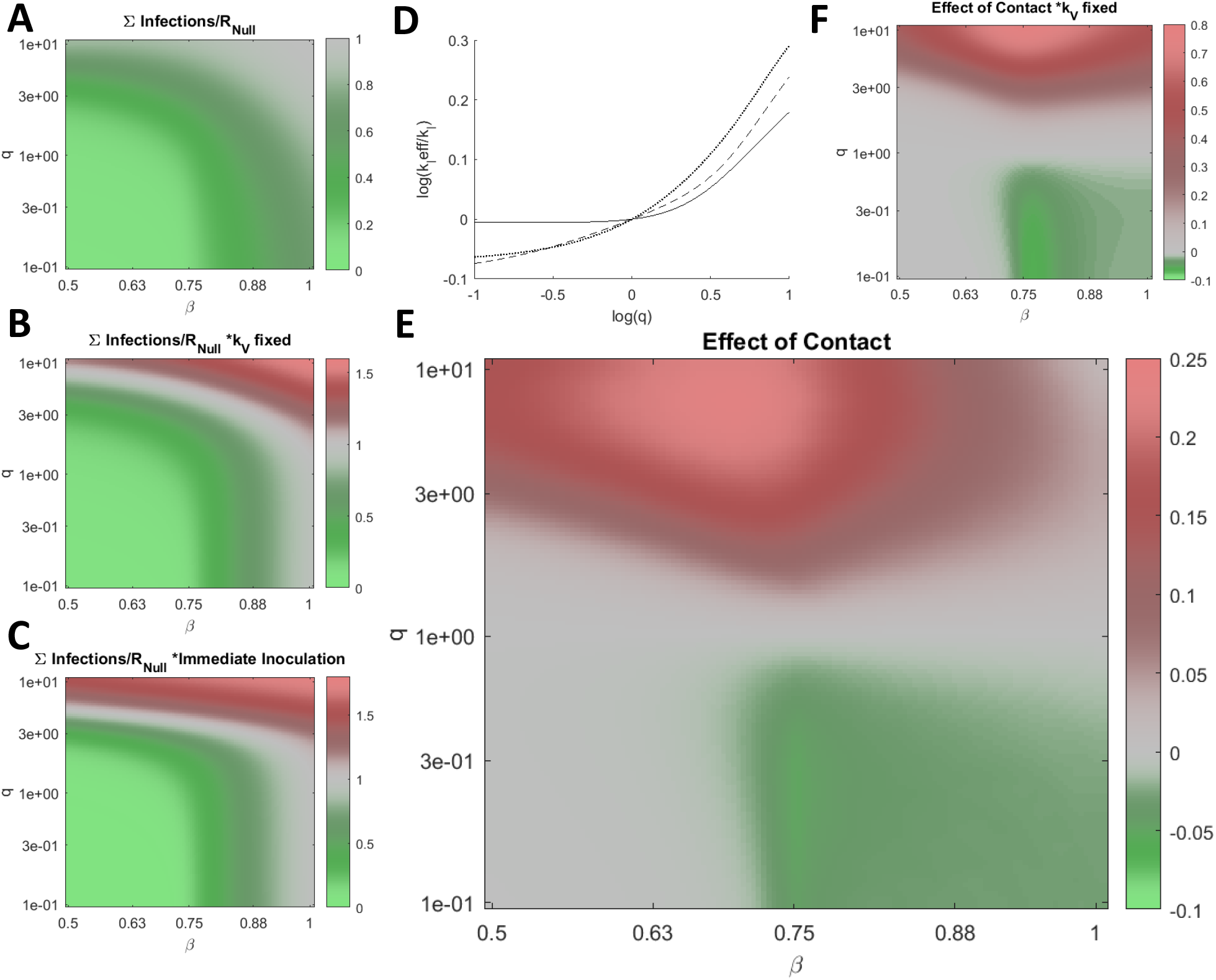
Post-vaccination contacts. **A**. The cumulative number of infections relative to no vaccination, *R*_*Null*_, for a range of *β* and *q*, optimizing *k*_*V*_ for each condition. **B**. Same as in A. for fixed *k*_*V*_=*0*.*03* representing the inoculation of 3% of Naïve hosts per day. **C**. Same as in A. for the case where all Naïve hosts are immediately inoculated. **D**. Log of the effective *k*_*I*_, *kIeff*, relative to *k*_*I*_=*0*.*2*, the benchmark, vs. log of *q* for *β=*[*0*.*5* (dotted line), *0*.*7* (dashed line), *1* (solid line)]. **E**. The cumulative number of infections relative to *R*_*Null*_ and *q=1* scaled by the fraction of infections due to vaccine escape, ∑Escape(q)/∑Infections(q)(∑Infections(q)-∑Infections(q=1))/R_Null_, for a range of *β* and *q*, optimizing *k*_*V*_ for each condition. **F**. Same as in E for fixed *k*_*V*_=*0*.*03*. **A-C/E/F**. 961 values were computed for each panel and 4x by 4x bilinearly interpolated points are displayed.

We additionally consider the cumulative infections added or subtracted relative to the outcome corresponding to *q=1* and scaled by the fraction of infections due to vaccine escape (Fig. 3E). Varying *q* within an order of magnitude alters the cumulative number of infections added or subtracted by more than 20% of the cumulative number of infections in the absence of vaccination, *R*_*Null*_, again demonstrating the critical role played by inoculated hosts with respect to vaccine escape. The landscape is similar when then rate of vaccination is fixed (Fig. 3F).

## Discussion

Epidemics can be mitigated through the reduction of host-host contact (quarantines and other similar measures) and vaccination. A reduction in contact carries non-negligible social and economic burdens so that, when vaccination becomes possible, the continuation of such interventions might appear unnecessarily costly. Formulating an optimal vaccination strategy to balance these pressures is challenging and is further complicated by the possibility of vaccine escape. Escape variants emergingas a result of vaccination are not expected to be more infectious than the originating strain, otherwise, such variants would likely already be in circulation prior to vaccination.

However, newly emergent variants are not always much less infectious and can be responsible for non-negligible disease incidence^17,18^. Here, we demonstrated that, even when escape variants are modestly less infectious, there exists an optimal vaccination distribution rate such that exceeding this rate increases the cumulative number of infections. This optimal rate depends on the infectivity of the escape variants. Such a prediction is impractical at the time of writing, that is, in the very first days of the vaccination campaign, and the cost of overestimation of the optimal vaccination rate is far less than that of underestimation. However, to our knowledge, this phenomenon, analogous to the evolution of antibiotic resistance, is not widely appreciated and, as such, seems to warrant discussion. Of more practical concern is the role of inoculated hosts in the emergence of escape variants. Within low-dose antibiotic regimes ^14,15^, hosts are susceptible to infection with the originating variant, and in such hosts, the pathogen is subjected to elevated selective pressures towards the emergence of resistance. Similarly, within inoculated hosts, the virus is subjected to gradually increasing selective pressures towards the emergence of resistance while the intra-host population remains sufficiently large to explore a substantial fraction of the mutation space. We demonstrated that moderately increasing or decreasing the host-host contact rates for inoculated hosts only can substantially aid or hinder the vaccination campaign. The time between vaccination and the acquisition of immunity can be readily approximated from clinical endpoints^19,20^ and is likely to be short enough that the societal costs of limiting post-vaccination contact would be outweighed by these benefits.

## Limitations

In this study, we leveraged classical modelling techniques to elucidate the factors that could substantially impact the outcome of any vaccination campaign. Although these results are broadly applicable, they are not necessarily instrumental for predicting quantitative outcomes for the current SARS-CoV-2 pandemic or any other particular epidemic. Similarly, the model presented in this study is not designed to forecast long-term outcome, a topic that has been thoroughly addressed for the case of SARS-Cov-2 and more generally^7-13,21-23^.

The results presented here appear to be of immediate interest in relation to the vaccination campaign against SARS-Cov-2 that is beginning at the time of this writing. However, although diversification of the virus is apparent^24-26^ and the potential for escape is being actively investigated^5,6,27,28^, it cannot be ruled out that all escape variants have a substantially reduced infectivity, represented by *β*<<*1*, in which case the highest possible vaccination rate will be optimal and the benefits specific to post-vaccination contact limitation could be negligible.

## Conclusions

Depending on the infectivity of escaping, vaccine-resistant mutants, the optimal vaccination rate with respect to the cumulative number of infections can be lower than the maximum rate. Contact rates for inoculated hosts can have a substantial impact on the outcomes of vaccination campaigns. Even a brief and moderate limitation of contacts in this well-defined population can potentially mitigate epidemics. These results might merit consideration for an immediate, practical public health response.

## Data Availability

No data is required for the reproduction of the results contained in this work.

## Author Contributions

NDR, YIW, and EVK conceived of the study; NDR constructed the model; NDR, YIW, and EVK analyzed data; NDR and EVK wrote the manuscript that was edited and approved by all authors.

## Acknowledgements

The authors thank Koonin group members for helpful discussions. NDR, YIW, and EVK are supported by the Intramural Research Program of the National Institutes of Health (National Library of Medicine).

